# Development of an interdisciplinary network to improve the capacity to conduct digital legacy research: a quality improvement initiative

**DOI:** 10.64898/2026.08.06.26359876

**Authors:** Amara Callistus Nwosu, Andrew Tibbles, Christian Goodwin, Lauren Kaye, Sarah Stanley

## Abstract

**Background:** Digital legacy (the digital information available about someone following their death) has increasing societal importance as personal assets and interactions become increasingly digitized. Healthcare professionals often have a limited understanding of how to address digital legacy in practice, and there is a lack of interdisciplinary networks to improve education, research, and professional development in digital legacy.

**Objective:** This paper describes the development of an interdisciplinary initiative designed to build research capacity and develop consensus-based recommendations for integrating digital legacy into palliative care.

**Method:** Over 12-months, we conducted interdisciplinary engagement activities with diverse stakeholders, including clinicians, designers, and sociologists. We used a modified World Café method to facilitate dialogue and capture feedback on how memories are digitally curated, the management of digital estates, and intergenerational perspectives on digital legacy.

**Results:** We identified eight core recommendations for research and policy, including promoting digital legacy education, supporting policy development, and broadening the scope of interdisciplinary research. Our discussions highlighted the complexity of modern digital estates and the need for legal and ethical frameworks to protect individual rights.

**Conclusions:** The Network demonstrates that interdisciplinary collaboratives can address important issues relating to digital legacy, which provides a foundation to conduct collaborative research that improves the management of digital legacies in society.

## BACKGROUND

A digital legacy is the digital information available about someone following their death.[1] A digital legacy is shaped by the interactions and information created during a person’s life, such as social media profiles, photos, videos, and gaming profiles.[2] Digital legacy is increasingly important societal issue (due to digitisation of most information associated with modern life[3]), with researchers and policymakers exploring how to best support society to manage digital assets after death.[4] It is likely that different types of digital legacy planning and support is needed for different age-cohorts, with younger people more likely to have a larger digital footprint in their lifetime compared to older people.[5, 6]

Previous research identifies three key priorities to improve societal awareness of digital legacy. Firstly, ‘digital memories’, describing the use of technology to create, curate, and archive memories (e.g. artificial intelligence (AI) generated content, which can be used to digitally recreate aspects of a deceased person).[7, 8] Secondly, the ‘digital estate’, describing practical issues of managing an individual’s digital assets. Digital estates may include assets with financial value (e.g., cryptocurrency) and sentimental value (e.g., social media accounts).[9] Thirdly, acknowledging ‘generational perspectives’ about digital legacy is important to consider the different attitudes and beliefs across generational cohorts regarding data ownership, privacy, and the conversation about managing digital assets after death.[5, 10]

Although the societal importance of digital legacy in increasing, healthcare professionals lack understanding about how to incorporate into advance care planning with patients and caregivers, when discussing their preferences of how their digital assets are managed after death.[7] Research is needed to improve digital legacy management; however, digital legacy is limited by a lack of collaborative work between researchers in the fields (e.g. computer science, law, sociology, design, and palliative care) needed to research the multi-faceted topic of managing digital post-mortem.[11] [12] Interdisciplinary approaches may be best suited for digital legacy research (compared to siloed and multidisciplinary approaches), as they integrate knowledge, methods, and theories from different fields to create a new, cohesive, and innovative whole, rather than just combining separate, independent contributions.[13, 14] However, there is a lack of established interdisciplinary networks in palliative care digital legacy research. Consequently, Cross-disciplinary research is needed to help global societies manage the growing volume of post-mortem digital data.[1] [15]

## AIM

The aim of this paper is to describe the development of an interdisciplinary initiative designed to build capacity for digital legacy research. Furthermore, this paper presents consensus-based recommendations developed by the network to improve the future delivery and integration of digital legacy research into palliative care practice.

## METHODS

Over 12-months, we conducted engagement activities to establish the ‘Digital Legacy, Design and Technology Network.’ We recruited stakeholders from different disciplines, including healthcare professionals, designers, philosophers, sociologists, computer scientists, and ethicists. We chose the approach of developing University community engagement activity to improve collaboration between stakeholders to address complex societal problems and identify recommendations for future digital legacy palliative care research.[16, 17]

Participants were invited using a convenience sampling approach consistent with methodologies employed in our previous digital health priority-setting research.[12] We solicited a convenience sample of professionals (working across the North West of the UK) working in palliative care, technology and design (including physicians, nurses, social workers, designers, academics, spiritual care staff, and managers) who were interested in palliative care research. To ensure diverse representation beyond our immediate academic and clinical circles, we advertised the network and upcoming meetings via social media platforms and targeted professional email lists.

### Interdisciplinary meetings

We conducted two interdisciplinary meetings to bring together diverse perspectives from design, palliative care, and digital sectors.

### Meeting 1: Scoping and introduction (September 2023)

The first meeting aimed to introduce major themes and interconnected areas of digital legacy within palliative care. The structure of the day consisted of morning presentations focusing on the current landscape of digital legacy, the role of design at the end of life, and the incorporation of digital legacy into current palliative care practice. The afternoon session involved workshops addressing inequalities in digital legacy, designing the future of palliative care, and the potential of new technologies.

### Meeting 2: Identifying practical applications of digital legacy and establishing recommendations for future work (March 2024)

In the second meeting we identified practical applications to improve digital legacy discussions in practice and we developed recommendations for future research and policy. The meeting involved presentations from multidisciplinary experts, including a photographer discussing applications of Artificial Intelligence (AI) in grief, a museum curator on technology-led dementia awareness, and a designer on palliative care innovation.

### Data Collection

We used a modified World Café Method to capture feedback.[18] We selected this method due to its ability to foster open dialogue and encourage the cross-pollination of ideas regarding complex digital legacy issues. The session was structured around three twenty-minute rounds of conversation, with participants rotating between three tables. Three facilitators (A.C.N, S.S, and A.T) moderated the tables, using open questions around three pre-defined themes: (1) the creation and curation of ‘digital memories’; (2) the practical management of ‘digital estates’; and (3) ‘intergenerational perspectives’ on digital legacy conversations.

Data collection was an iterative process; during each round, facilitators and table scribes recorded stories and made detailed written notes of the dialogue. To ensure continuity, the facilitators summarized the core content for each incoming group, allowing new participants to build upon previous discussions. The activity concluded with a summary meeting, where the main notes were shared with the wider group for immediate reflection. Following the event, we summarised the feedback and shared the summary with all attendees (by email) and we encouraged participants to respond with further comments prior to development of the final recommendations.

## Results

### Participant Demographics

The first meeting (September 2023) was attended by 23 participants, and the second meeting (March 2024) by 27 participants. Attendees represented different disciplines, including healthcare clinicians (physicians and nurses), healthcare researchers, design and arts researchers, museum directors, sociologists, counsellors, business analysts, and Patient and Public Involvement (PPI) representatives (Table 1).

**Table 1:** Summary of attendees of the two Digital Legacy Technology and Design Networking Meetings.

|  | **Meeting 1**<br>**n 23 – Meeting 1st**<br>**September 2023** | **Meeting 2**<br>**N = 27**<br>**2<sup>nd</sup> March 2024** |
| --- | --- | --- |
|  | **N (%)** | **N (%)** |
| **Healthcare professionals** | **3** | **4** |
| **RESEARCHERS** |  |  |
| **Healthcare researchers** | **4** | **8** |
| **Sociology researcher** | **1** | **1** |
| **Design researcher** | **5** | **1** |
| **Art and design researcher** | **1** | **1** |
| **Museum director** | **1** | **1** |
| **Counsellors** | **0** | **1** |
| **PPI representative** | **1** | **1** |
| **Business analyst** | **1** | **0** |
| **MANAGERS** |  |  |
| **Research managers** | **5** | **2** |
| **Healthcare manager** | **0** | **6** |
| **Healthcare educator** | **0** | **1** |
| **Academic editor** | **1** | **0** |

### Outcomes of the group discussions

The following section describes details from in-depth discussions of the three themes which arose during the World Café discussions:

#### Curation of Digital Memories

Participants identified several digital formats that constitute modern memory making, ranging from recording digital media (video and audio), social media, virtual reality (VR) and avatars (e.g. image manipulation and voice cloning). Attendees spoke about how physical and digital artefacts produce different memory-making experiences for the user. For example, physical items can be manipulated physically and may induce sentimentality and feeling of exclusivity for the recipient or owner. Comparatively, digital assets provide opportunities for electronic storage and archiving, digital manipulation, reproduction and dissemination. Participants spoke of specific challenges associated with digital assets, such as issues with accessing data, ownership, privacy and authenticity of data (especially due to the ability to manipulate digital information).

#### Management of the Digital Estate

Attendees catalogued the complexity of the modern digital estate, which includes financial assets (crypto, banking), subscription services, and cloud storage. Key barriers identified included large volumes of data, password management, and a lack of knowledge regarding legal regulations. Facilitators for discussion included open conversations, the use of password managers, and human centred design of technological systems.

#### Intergenerational Perspectives

Participants noted that while all generations demonstrate a desire to preserve memories, their approaches differ. Younger generations were characterized as ‘super users’ who were digital natives and more comfortable with using social media and with AI. Older generations may struggle with multiple platforms and rely on younger family members for technical support. A shared concern across cohorts was the safeguarding of data and the concern that the public lack support, guidance and knowledge of how to manage their digital assets.

#### Recommendations

We used the group discussions to generate eight recommendations for digital legacy research, policy, and practice. These are summarised below and further information about the recommendations are presented in Table 2.

1. Promote digital legacy education
2. Support development of digital legacy policy
3. Improve awareness of cybersecurity for digital legacy
4. Broaden scope of digital legacy research
5. Develop stakeholder partnerships with skills in digital legacy
6. Improve awareness of the digital legacy rights of Individuals
7. Develop legal and ethical frameworks for the curation, use, management and deletion of digital assets
8. Improving Implementation of digital legacy best practice.

**Table 2:**
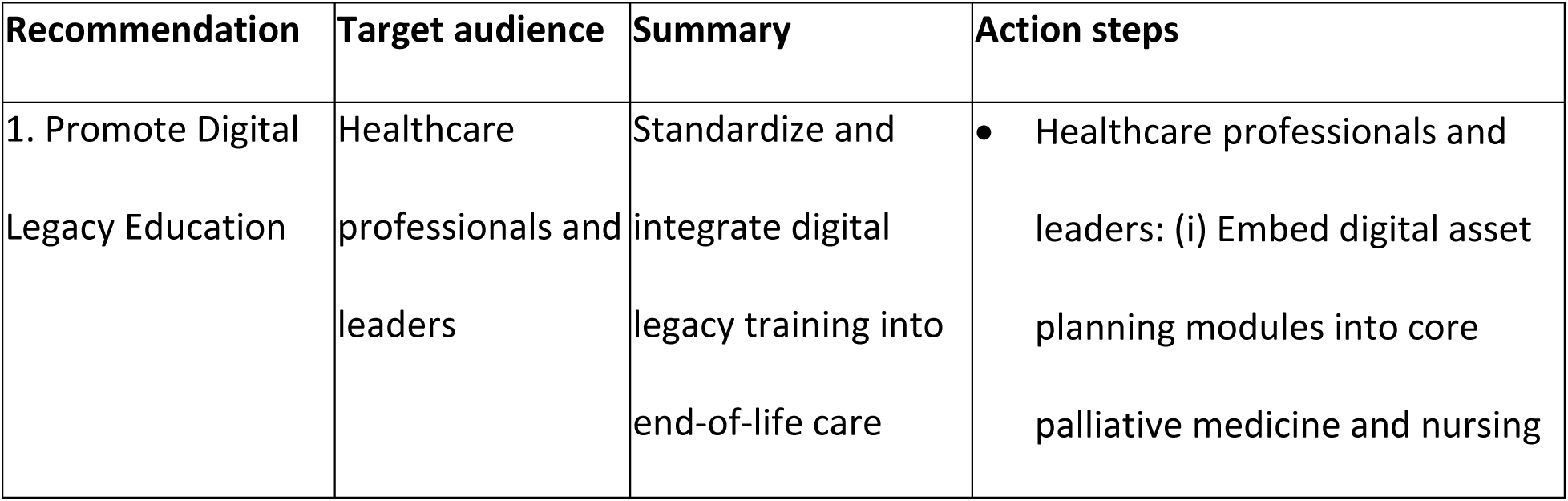
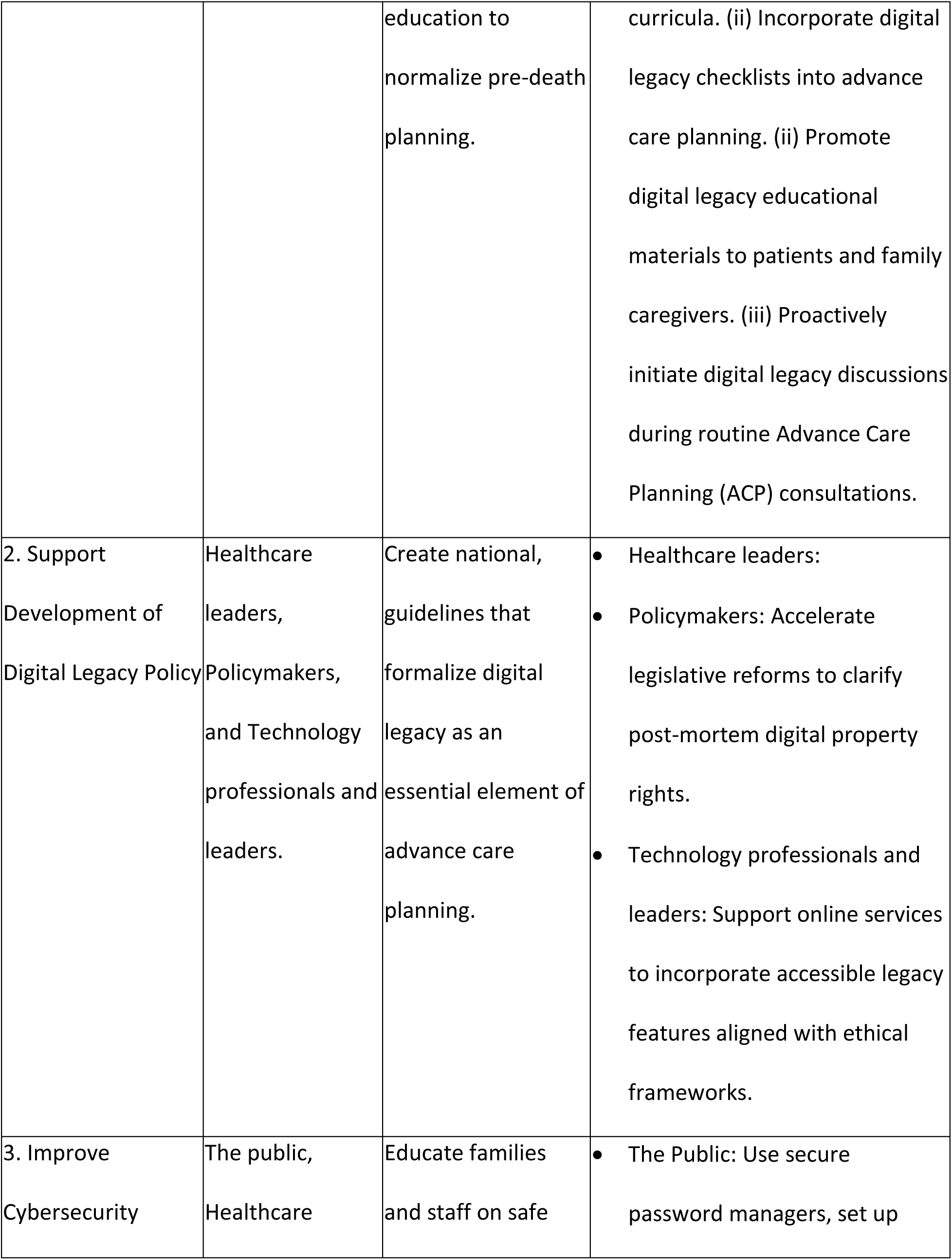
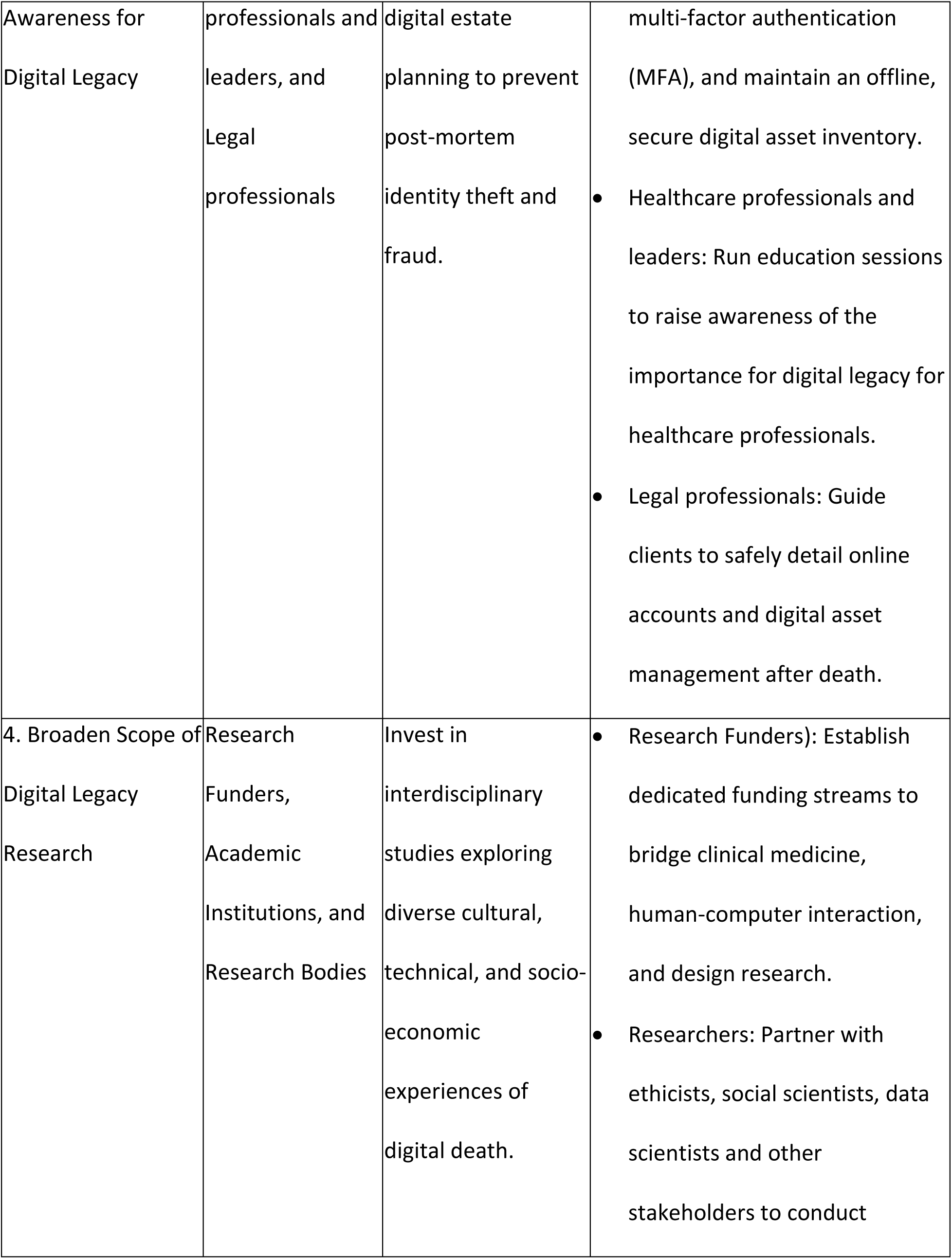
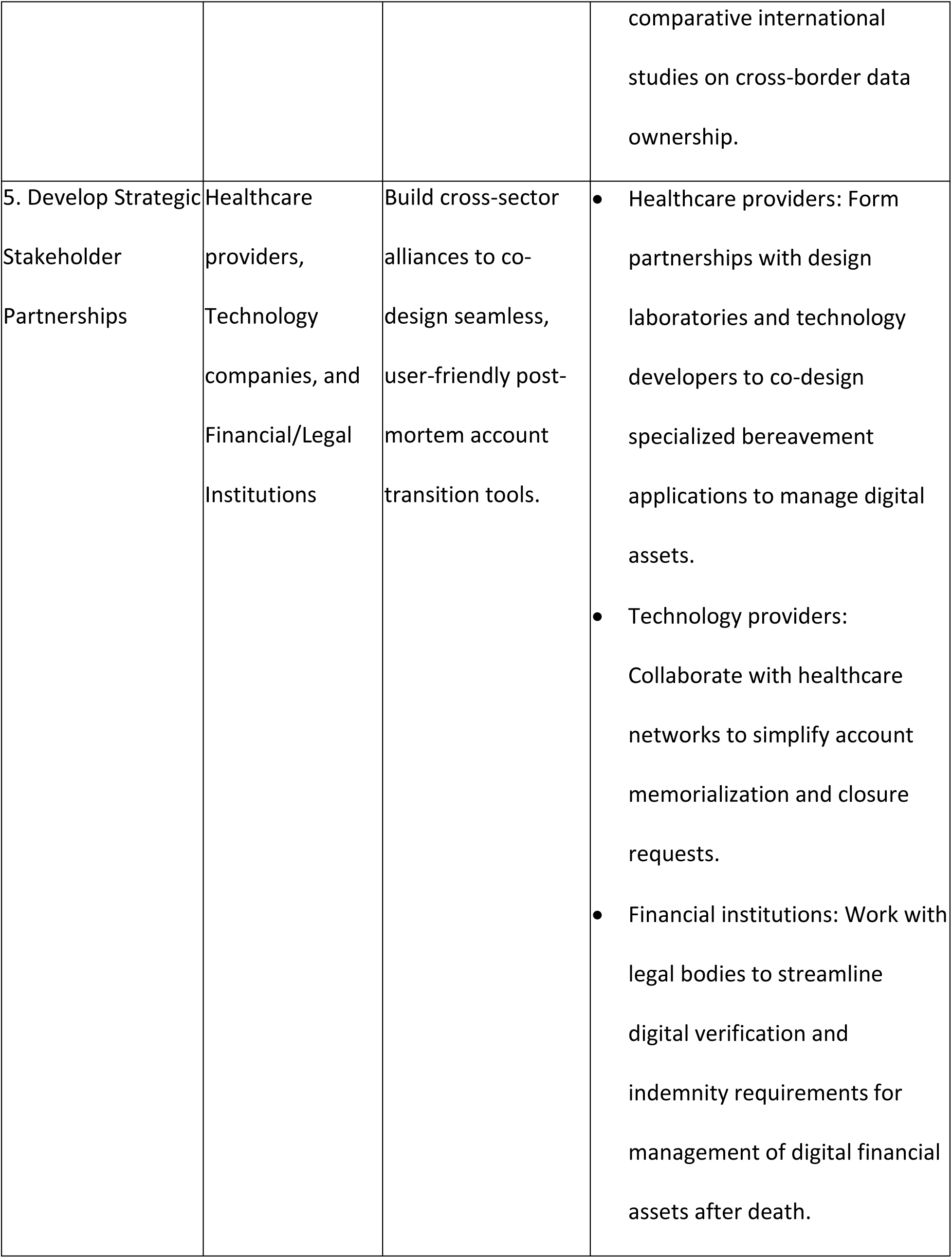
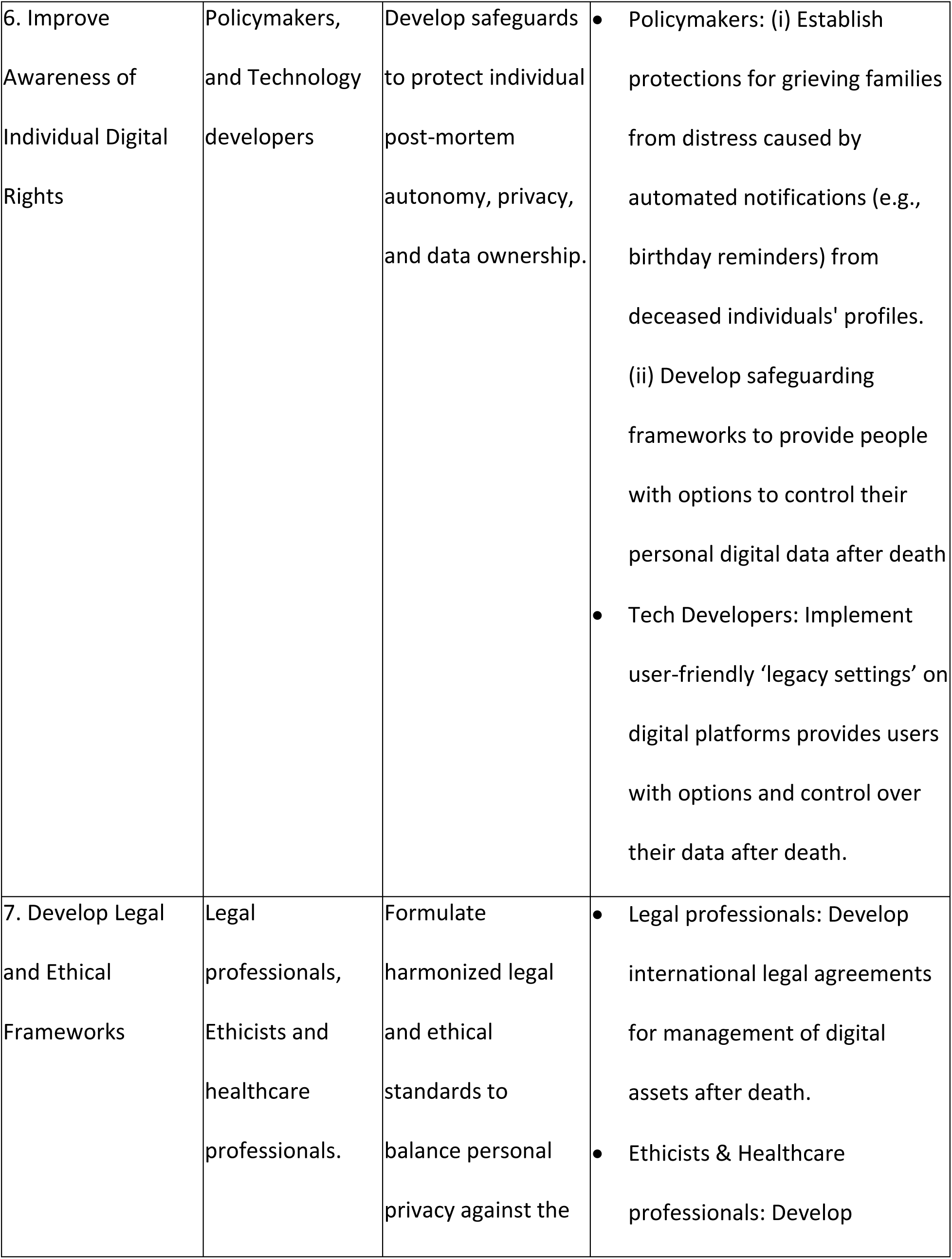
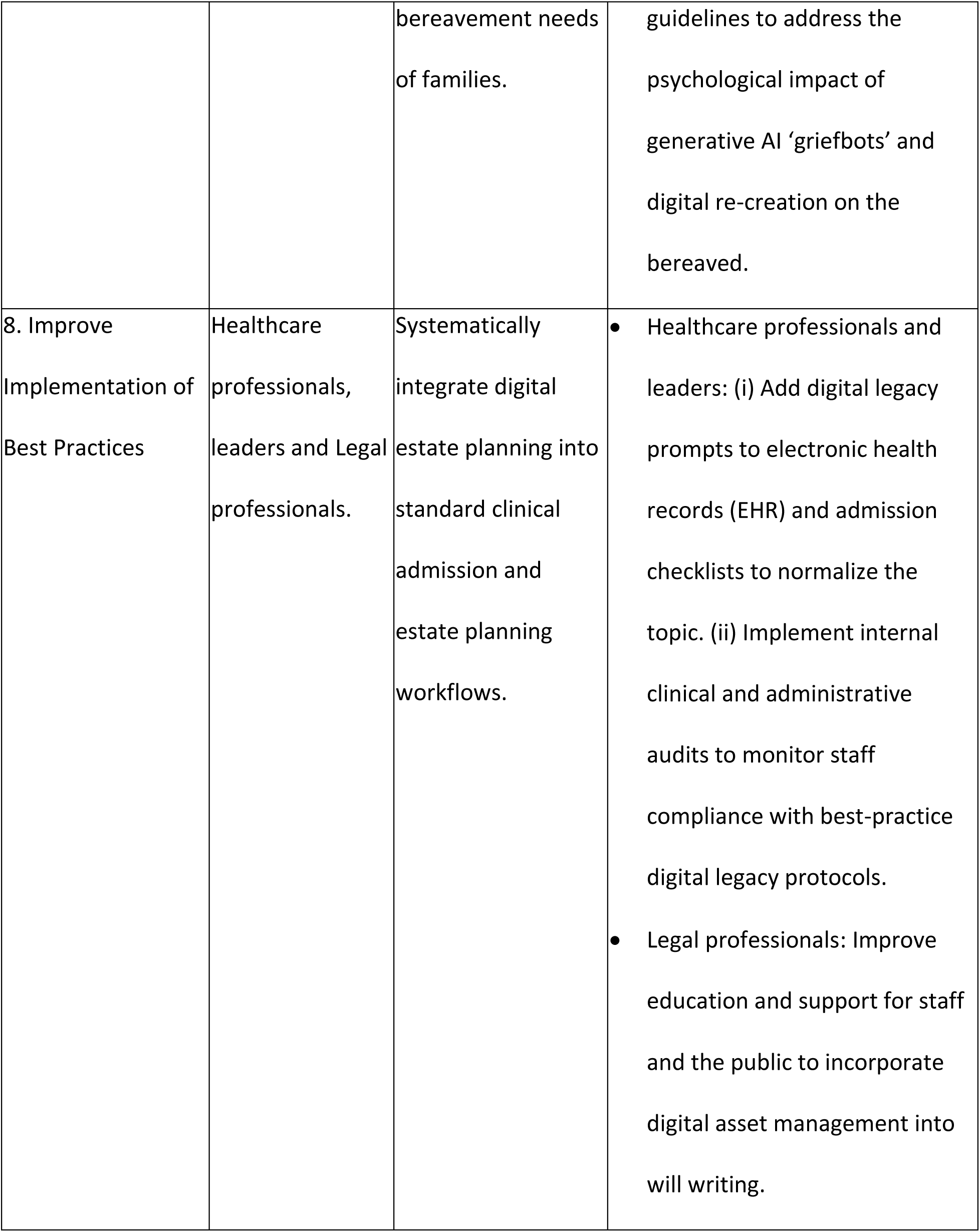
Recommendations to facilitate collaborative Digital Legacy Research.

## Discussion

### Summary of the main findings

Our paper demonstrated that we successfully developed a network connect a diverse group of individuals and organisations to provide insight and discussion into how technologies can be used to create, curate, and archive memories. The outcomes of the group included the development of an interdisciplinary network to support research, and a developed list of recommendations for future digital legacy research.

### Strengths and new knowledge

The paper is unique in reporting on the development of an interdisciplinary network to provide a foundation for digital legacy research. Our paper provides a detailed overview of the process, meaning that the procedure should be reproducible to others who wish to create similar networks with stakeholders in their regions.

### Relationship to previous research

This paper highlights opportunities to enhance digital legacy palliative care research capacity through collaborative opportunities across clinical and research settings. Our findings support previous research which identifies the benefit of interdisciplinary collaborations for research in palliative care (e.g. conducting research, successful research grant submissions and building research capacity).[16] Our recommendation to improve education for palliative care healthcare professionals supports previous work reporting how healthcare professionals feel unprepared to deliver digital legacy conversations with patients.[7, 19] Also, our work is consistent with studies which advocate for integrating digital legacy into advance care planning (ACP), which is supported by organizations like the Digital Legacy Association.[2, 20] Our recommendations about the importance of legal and policy frameworks are consistent with international efforts to develop laws about recognizing digital assets as personal property.[21] [22]

Our findings support existing arguments from diverse stakeholders, that international agreement is needed to improve regulation of how digital assets are management before and after death.[23] Consequently, our work supports research describing the need for interdisciplinary approaches to improve public facing digital platforms.[24] These approaches may involve development of policy to improve design and oversight of digital platforms, through cybersecurity initiatives password managers, multi-factor authentication, and platform-specific tools like Apple’s ‘Legacy Contact’).[25, 26]

While there is general agreement on the need for planning, certain recommendations face significant legal, ethical, and corporate opposition. Firstly, many digital service providers use ‘no right of survivorship’ clauses in their Terms of Service, meaning content licenses (e.g. for eBooks or music) expire upon death, which conflicts with ensuring individuals and caregivers retain ownership of their data.[27, 28] International data protection laws may create barriers for people to access to digital data of the dead (for example, the EU and UK General Data Protection Regulation (GDPR) have no specific policy for the dead). Further, AI use to manipulate data of deceased people introduces a range of practical, privacy and ethical challenges, particularly during grief and bereavement.[29, 30] [31]

Our paper highlights the barriers to conducting interdisciplinary research, including issues like governance issues, lack of awareness and skills within research groups, and a lack of cross-sectional leadership and policy in digital legacy research. Our work is consistent with other studies which describe these challenges specific to palliative care research partnerships,[32] and general interdisciplinary research.[33]

Our study proposes eight recommendations to improve digital legacy research which is consistent with the finds of previous. [5] Our network support previous evidence that shows how interdisciplinary technology projects succeed with involvement of people who can bridge the gap between technical systems architecture and social theory and encourage real world practical translation of ideas, alongside the theoretical work.[34] Our findings are also aligned with work to reduce digital inequalities in palliative care, [20, 35, 36] by describing the benefits of diverse digital stakeholders working together to research palliative care digital health.[2, 12, 24, 37]

### Limitations

Our work describes a regionally focused network of stakeholders in the north of England, meaning the findings of this work may not be translated to other settings. Although the predefined themes for the world café discussion were chosen to ensure that attendees spoke about the specific areas of interest, there is potential pre-defining limited the breadth of the discussion and increased the risk of facilitator bias (i.e. participants more likely to discuss topics of interest to the facilitator rather than discuss their own views). However, we tried to reduce the risk of facilitator bias by using open questions during the discussion, encouraging attendees to expand their answers, and provide new insights during both the small group and wider group sessions. Although the network included a diverse mix of participants there were unrepresented areas (e.g. ethicists, data scientists and members of the public) meaning we were unable to obtain viewpoints from these groups.

### Implications for policy, practice and research

The outcomes of our study can be used by relevant stakeholders to improve interdisciplinary working in digital legacy to focus on improving collaboration and culture, capacity building and developing clear guidance on research processes and procedures to reduce friction points for stakeholder collaboration. Future research should investigate how digital legacy needs vary across global and non-Western cultural contexts to address the regional limitations of current studies. Furthermore, interdisciplinary researchers should explore how to integrate underrepresented experts, such as ethicists and data scientists, into research frameworks. This work should expand beyond clinical settings to examine the role of schools, the arts, and the media to normalize conversations about digital legacy and how to improve digital legacy literacy in society. As technology evolves, research is required to understand how developments in new technologies (e.g. such as quantum computing) will transform the long-term security, encryption, and preservation of digital estates.[26] Researchers should use participant-led methods to reduce risk of facilitator bias and uncover emerging themes that predefined frameworks may overlook. Finally, researchers should aim to identify strategies that reduce digital inequality, to ensure marginalized groups have agency of their digital assets, whilst examining the long-term psychosocial, ethical and legal issues associated with the management of digital legacy in the modern world.

## Conclusion

The ‘Digital Legacy, Design and Technology Network’ has demonstrated that an interdisciplinary model can successfully bridge gaps between fragmented fields like design, palliative care, and technology. By coordinating a diverse cross-section of stakeholders (including clinicians, researchers, and creative professionals), this initiative has provided a framework to improve research capacity and professional awareness. Future interdisciplinary digital legacy research should aim to identify how emerging technologies can be best used to empower individuals, support families, and ensure that the management of digital legacies is integrated into holistic, ethical, and equitable end-of-life care for all of society.

## Author Contributions

### CRediT Author Contributions

- **Amara Callistus Nwosu (ACN):** Conceptualization, Methodology, Funding Acquisition, Investigation, Project Administration, Supervision, Writing – Original Draft, Writing – Review & Editing.
- **Andrew Tibbles (AT):** Investigation, Formal Analysis, Writing – Review & Editing.
- **Christian Goodwin (CG):** Formal Analysis, Writing – Review & Editing.
- **Lauren Kaye (LK):** Writing – Review & Editing.
- **Sarah Stanley (SS):** Conceptualization, Methodology, Investigation, Project Administration, Supervision, Writing – Original Draft, Writing – Review & Editing.

## Statements and Declarations

### Ethical considerations

This project was classified as a service-evaluation and quality-improvement project and did not constitute clinical research. Therefore, formal institutional ethics committee approval was not required.

### Consent to participate

Informed consent to participate was obtained verbally from all stakeholder participants prior to the commencement of the network meetings and modified World Café workshops.

### Consent for publication

Not applicable. No individual personal data, images, videos, or highly specific identifying details of participants are included within this manuscript.

### Data availability

The qualitative datasets (raw workshop notes and scribed reports) generated and analyzed during the current study are available in the in supplementary material. Anonymized summaries of the thematic discussions may be made available from the corresponding author upon reasonable request.

## Supplementary files

Supplementary file 1: Outline agenda for the meetings

Supplementary file 2: Summary of networks used to invite people to the digital legacy deign and technology network

Supplementary file 3: Raw data from workshops

