## Supplementary file 1 for "Development of an interdisciplinary network to improve the capacity to conduct digital legacy research: a quality improvement initiative"

Meeting agendas – Digital legacy, Design and Technology meetings

**Agenda for meeting 1 - 20th September 2023**

| Time | Topic | Presenter |
|---|---|---|
| 0930 - 1000 | Registration | |
| 1000 - 1020 | Introduction to the day | Dr Amara Callistus Nwosu Senior Clinical Lecturer in Palliative Care, Lancaster University |
| 1020 - 1100 | An overview of digital legacy, the 'Digital Legacy Association'<br>and 'My Wishes | James Norris (Founder and European Lead of the Digital Legacy Association and My Wishes) |
| 1100 - 1120 | Tea, coffee and networking discussion | |
| 1120 - 1200 | Designing meaning at the end of life. | Dr Farnaz Nickpour (Reader in Inclusive Design & Human-Centred Innovation at The University of Liverpool, and Director of The Inclusionaries Lab for Inclusive & Human Centred Design Research). |
| 1200 - 1230 | Preparing for digital legacy in Palliative Care. | Sarah Stanley (PhD Nurse Researcher, Marie Curie Hospice Liverpool). |
| 1230 - 1330 | Lunch | |
| 1330 - 1415 | Inequalities in digital legacy (workshop). | Christian Goodwin (Fulbright USA-UK Scholar and medical student, University of North Carolina School of Medicine, North Carolina, USA). |
| 1415- 1500: | Designing the future in digital legacy (workshop). | Andrew Tibbles (PhD design student, the University of Liverpool). |
| 1500 - 1515: | Break, tea, coffee, networking. | |
| 1515 - 1600 | New technologies and digital legacy (workshop). | Professor Mark Taubert (Consultant in Palliative Care at Velindre University NHS Trust and Professor at Cardiff University School of Medicine. |

| | |
|---|---|
| 1600 - 1605: | Summary and close. |

**Meeting 2 – 20th March 2024**

| Time | Topic | Presenter |
|---|---|---|
| 0930 - 1000 | Registration | |
| 1000 - 1030 | The Digital Afterlife of Grief | Ginger Liu - CEO/Founder at Ginger Media & Entertainment/PhD Researcher(MFA in Photography and a BA (Hons)in Contemporary Media Practice) |
| 1030 - 1100 | House of Memories: museum-led dementia awareness programme using technology | Dawn Carroll (Liverpool Musuems) |
| 1115 - 1145 | Designing the future of palliative care: the 'Designer in Residence' programme | Andrew Tibbles (PhD design student, the University of Liverpool) |
| 1145 - 1200 | Artificial intelligence in palliative care: opportunities & challenges | Dr Amara Callistus Nwosu Senior Clinical Lecturer in Palliative Care, Lancaster University |
| 1200 - 1230 | A young person's perspective on digital legacy: an interview with a 16 year old | Sarah Stanley (PhD Nurse Researcher, Marie Curie Hospice Liverpool). |
| 1230 - 1330 | Lunch | |
| 1330 - 1500 | Workshops<br>Attendees will rotate around 3 tables to discuss practical applications of the following topics:(1) Digital memories,(2) Digital estate and (3) Opening up the conversation across generations. | Amara Nwosu<br>Sarah Stanley |
| 1500 - 1520 | Tea, coffee and networking | |
| 1520 - 1550 | Feedback from workshops | |

| | |
|---|---|
| 1550 - 1600 | Summary and close |
| 1500 - 1515: | Break, tea, coffee, networking. |
