## Supplementary file 2 for "Development of an interdisciplinary network to improve the capacity to conduct digital legacy research: a quality improvement initiative"

Appendices – summary of networks used to invite people to the digital legacy deign and technology network

The study invitation was distributed electronically to an estimated 13292 individuals through the following networks and methods:

- **Professional Networks:**
  - **International Collaborative for Best Care of the Dying Person:** A multidisciplinary international research group (n=79).
  - **CHAIN (Contact, Help, Advice and Information Network):** The UK National Health Service (NHS) technology interest email list (n=598).
  - **Marie Curie Hospices UK:** Email distribution to all staff across nine hospices (estimated reach n=450).
  - **North West Technology and palliative care interest group:** Including leasers from the NIHR North West Regional Research Delivery Network (RRDN) and professionals who have previously attended regional palliative care digital health events and provided consent to be contacted by email (estimated reach n=50).
- **Social Media:**
  - **X:** Four "posts" were sent from the lead author's profile using palliative care hashtags (e.g., #hapc, #hpm, #palliativecare) individuals.
  - LinkedIN
  - (Reaching approximately n=12,058)
- **Direct Outreach:**
  - **Targeted Emails:** Sent to 57 professionals from a previous palliative care technology event who had consented to receive information about future studies.
