## Supplementary file 3 for "Development of an interdisciplinary network to improve the capacity to conduct digital legacy research: a quality improvement initiative"

Raw data from workshops

**Table 1: Digital memories**

- What examples of digital memories are you aware of?

Voice Notes

Digital windows

Photos/videos we upload online: digital frames/Facebook memories

Emails/Messages

Virtual reality 'recreation of a memory'

Dating sites

Deep fake

Old Facebook posts

Music playlists 'your soundtrack'

Memorial websites

Google

Bank transactions

Data tracking

Apps

Online interaction

Cookies

Cloud memory

Podcasts

- What are the differences between digital and physical memories?

When is it digital? Does this include hard drive/floppy disk?

**Physical**

- Control, ownership and exclusivity of physical objects
- More sensory: touch and smell
- Tangible

- Value in being irreplaceable
- Fewer physical items increase value, compared to lots of digital items decrease value
- Muscle memory – physical interaction

- How may people feel about receiving digital memories?
- Overwhelmed with responsibility (especially without information on what to do!)
- Distressed (context/time dependant. Historical different to personal)

- How do you feel about the digital memories you own?

  - What are the opportunities?
  - Immortal quality of content
  - Foresight to how you want to be remembered
  - Curation control

  - What are the challenges?
  - Accessibility
  - Amount and Volume
  - Ownership
  - Audience
  - Historical warping modern perception
  - Accuracy

- How can we use what we have discussed today to plan how we make, manage and use digital memories?

<u>Make</u>

- Intentional
- Making unconscious creations = Precious

<u>Manage</u>

- Audiences
- Personal, close to you, public

<u>Use</u>

- Accuracy as desirable perception
- Curation vs Generation
- ‘Cleaning the house’ as a process of grief
- Editorial vs candid photos and the different values

**<u>Facilitator’s thoughts</u>**

*‘Digital legacy has the funeral problem ‘Who is it for?’ The goal of creation of our data is currently to earn a profit off the use which is not compatible with digital memories – so it won’t be sorted for those purposes. You are a consumer not a human.’*

**Table 2: Digital Estate**

- What examples of digital estate are you aware of?

- Monzo
- Go Henry
- Revolute
- CHASE
- Note apps (Evernote, notions, upnote, google)
- Google maps
- Fertility apps
- Birth records
- Wishes
- Medical records (NHS/COVID)
- Netflix/Disney+
- Stocks and shares
- Crypto currency
- Cloud storage (Google drive, iCoud, Dropbox, OneDrive)
- Online Banking
- Sun life insurance
- Social media
- Pension
- Apple pay/google pay
- Amazon
- Loyalty cards (Tesco, Nectar, M&S, Boots) = Reward points & currency
- PayPal
- Digital badges
- Gaming profiles
- Music profiles (Spotify)
- Subscription services
- Health apps (Strava, Google fit)
- Google (photos/documents)
- Work: Microsoft/OneDrive
- Klarna

- What are the facilitators and barriers in managing a digital estate?

Barriers

- Privacy
- So much data
- Pace of change
- Stolen phone or device
- Awareness of digital estate
- Laws/regulations
- Inequalities
- Passwords (and remembering passwords)
- What does happen when we stop paying (subscription)

Facilitators

- Age: Young & old
- Knowledge and skill
- Access
- Affordability
- Cost
- Privacy
- Organisation

- Conversations
- Access
- Password manager
- Legal
- Biometrics

- How do you feel about your own digital estate?

- What are the opportunities?
- Creating digital memory box
- Purposeful curation of memories
- Need to be active in creating memories
- Access
- Ease
- Shared information and accounts
- Interoperability (improved health by sharing data or using information to inform care)
- Environmental benefits
- Social inclusion
- Better understand digital estate
- Swedish death clearing???
- Being nice to people you leave behind

- What are the challenges?
- Online banking: no physical location. High street banks disappearing
- Music playlists: How would this be shared?
- Subscription services: What happens to data at the end of life? What are the legal implications?
- Clearing out digital house
- Organising digital life
- Social isolation
- Language

- How can we use what we have discussed today to help people to better manage their digital estate?
  - Where does digital legacy fit into power of attorney?
  - Digital will (increases awareness)
  - Discuss plan for EOL when setting up accounts/subscriptions.
  - Policy
  - Online/digital first
  - Digital services to manage

**Table 3: Opening up the conversation across generations**

- How do different generations talk and think about digital legacy?

**<u>What is similar?</u>**

- There is a desire to keep memories: sentimental value in physical and digital.
- Both digital and physical organisation is overwhelming, but digital is easier to 'just delete'.
- Essence of the person is not always captured in digital photos.
- Worry about losing 'genuine photos (eg looking away etc, particularly with some new phone features)
- 'Live' photos could capture essence.
- Phone/device IS a physical asset.
- Making memories easy to access should be important across generations.
- Conversations not currently happening.
- Safeguarding issues around passing on passwords.
- Don't always recognise we are using tech/creating legacy (Google/Alexa)

**<u>What is different?</u>**

- Shouldn't generalise: Some are tech savvy, some aren't.
- Different generations put different importance on DL.
- Legacy depends on the person.

**Younger generation**

- Big ideas
- Use AI day to day.
- Generate larger volume of digital data.
- Digital legacy is just legacy.
- Carefully curate social media
- See negative memories as important.
- 'Super users' of tech
- Will they want to deal with so many physical assets in the future?
- Seem to find digital more accessible.
- Sometimes like 'vintage' (eg Vinyl)
- Socials: Snap/BeReal

**Older generation**

- Sometimes find it difficult to use a number of different apps/technologies.
- Don't talk about digital legacy.
- Rely on younger generation for tech support.
- Don't think of DL as a thing
- Concern of data sharing (seems more acceptable for younger)
- Socials: Facebook/Instagram

- How can different generations learn, and help, each other to talk about digital legacy?
- Younger people could help older people to make digital memories.
- Objects mean nothing without a story behind it.
- Be brave to have discussions in all places (home, school, work etc)
- Losing control of accounts: hacking and messages in bereavement

- Digital wills might become the norm: Digital legacy should be incorporated.
- Potential to change digital memories to physical (reduce clutter, but risk of losing access)
- Younger people use physical memorials (e.g. covid/Manchester) but then share digitally.

- How can we use what we have discussed today to continue these discussions with more people, younger and older?
- Digital legacy should be considered over time rather than at end of life.
- Opportunities for training across generations (potential for chance in education)
- Encourage children to think about memory boxes: would be good for thinking of own mortality.
- Stories for life – Voice recording stories (make it less about when we die and more about the memories)
- Greater awareness: Make DL a social issue, government campaign?
- Concerns around younger generations becoming frustrated in workplaces such as NHS which are behind in tech world.
- Clinicians seem to object more to tech than patients!
- Children don't think about the future.
- Needs to be a normal conversation.

Inequalities

Overview from inequalities in digital legacy. Main points:

1) In healthcare and lay media," digital legacy" mostly referred to pragmatic elements of estate planning (e.g. providing next of kin access to online accounts and social media), rather than describing processes for preserving memories and subjective narratives.

(2) There is little published evidence about inequalities in digital legacy.

(3) Most authors writing about digital legacy and inequality are social scientists or computer scientists.

(4) Most resources are behind academic paywalls.

Questions to the group:

What questions do you have after this (brief introduction)

What questions surrounding DL and inequalities do you feel your discipline is best suited to address?

What research questions (as a group) try to answer in the next year? 5 years?

Where should we have these conversations? Who else should be included in these conversations?

Responses:

- Is DL important to the public? Does it matter? Do people understand how much of their life is digital?
- Risk of creating new inequalities
- Can tools be adopted for people who are frail? Use of proxies?
- What inequalities exist? What is the scale?
- How can we support equality in ACP?
- How can we incorporate DL into palliative care practices?
- How do we empower the individual?
- How do we make digital and data an accessible medium for creativity?
- Stakeholders: legal, funeral, educators, policy makers, HCPs

New technologies

Overview re: new technologies

1. Increased internet use (eg: for finding information)
2. Increased content creation (eg: Kate Granger, deathbed live)
3. Use of technologies such as VR and AR

Questions to the group:

How can we use new technologies help us to manage and create our own digital legacy in the future?

How can societies archive and preserve history and traditions in a digital age?

What are challenges are associated with new technology and digital legacy?

Responses

- Web
- Apps
- Cost of servers
- Reliability
- Financing/executing the contract
- Manage access/directions
- Need to make sure new technologies are accessible to your nominated person
- Public archives/link with current historical sites, eg: family tree (to futureproof)
- Money/cost involved for digital storage – Exclusionary?
- Ever increasing volume of digital assets – need to curate
- AI might be able to help
- Visiting virtual formats to futureproof the legacies. Avoiding obsolete tech such as CDs/videos
- Demystifying new tech: take work out of investigating them to make choice easier
- Using tech to create voice/life stories: could be AI to capture over time
- A programme to capture and organise digital assets: capturing access issues
- Who is DL for? Dying person or bereaved?
- Cleansing data: making decisions on what to keep
- Consent for making wishes known
- Issues in society: How do we manage these?
